# Lifespan analysis of repeat expression reveals age-dependent upregulation of HERV-K in the neurotypical human brain

**DOI:** 10.1101/2024.05.17.24307184

**Authors:** Taylor A. Evans, Arthur S. Feltrin, Kynon J. Benjamin, Tarun Katipalli, Thomas Hyde, Joel E. Kleinman, Daniel R. Weinberger, Apua C. Paquola, Jennifer A. Erwin

**Affiliations:** Lieber Institute for Brain Development, Baltimore, MD, United States of America; Johns Hopkins University School of Medicine, Department of Neuroscience, Baltimore, MD, United States of America; Johns Hopkins University School of Medicine, Department of Neurology, Baltimore, MD, United States of America; Johns Hopkins University School of Medicine, Department of Psychiatry & Behavioral Sciences, Baltimore, MD, United States of America; Johns Hopkins University School of Medicine, McKusick-Nathans Department of Genetic Medicine, Baltimore, MD, United States of America

**Keywords:** HERV-K, repeat biology, retroelements, aging, neurodegeneration, RNA-seq, Co-Expression

## Abstract

DNA repetitive sequences (or repeats) comprise over 50% of the human genome and have a crucial regulatory role, specifically regulating transcription machinery. The human brain is the tissue with the highest detectable repeat expression and dysregulations on the repeat activity are related to several neurological and neurodegenerative disorders, as repeat-derived products can stimulate a pro-inflammatory response. Even so, it is unclear how repeat expression acts on the aging neurotypical brain. Here, we leverage a large postmortem transcriptome cohort spanning the human lifespan to assess global repeat expression in the neurotypical brain. We identified 21,696 differentially expressed repeats (DERs) that varied across seven age bins (Prenatal; 0-15; 16-29; 30-39; 40-49; 50-59; 60+) across the caudate nucleus (n=271), dorsolateral prefrontal cortex (n=304), and hippocampus (n=310). Interestingly, we found that long interspersed nuclear elements and long terminal repeats (LTRs) DERs were the most abundant repeat families when comparing infants to early adolescence (0-15) with older adults (60+). Of these differentially regulated LTRs, we identified 17 shared across all brain regions, including increased expression of HERV-K-int in older adult brains (60+). Co-expression analysis from each of the three brain regions also showed repeats from the HERV subfamily were intramodular hubs in its subnetworks. While we do not observe a strong global relationship between repeat expression and age, we identified HERV-K as a repeat signature associated with the aging neurotypical brain. Our study is the first global assessment of repeat expression in the neurotypical brain.

## Introduction

Present in multiple copies, repetitive sequences, herein referred to as repeats, are a broad category of DNA sequences known to play crucial homeostatic roles within organisms including plants, insects, and humans through evolution (Ding et al., 2017; Petersen et al., 2019). In humans, repeats comprise over 50% of the human genome and have evolved to contribute to cell biology through more than insertional mutagenesis. However, following their discovery and causative link to hemophilia A, repeats have primarily been associated with their mutagenic capacity and disease (Gorbunova et al., 2021). A subcategory of repeats, retroelements, can move through an RNA intermediate, yet most of these elements are immobilized. Even when immobilized, repeats can contribute to the transcriptome of a cell, tissue, and organ (Schrader & Schmitz, 2018; Yamamoto et al., 2022).

Expression of repeat sequences poses a threat to genomic stability but is particularly relevant in the healthy human brain as it is the tissue with the highest detectable repeat expression (Bogu et al., 2019). Given the functional consequences of repeat expression in both healthy and diseased tissue, repeat sequences are regulated at the genomic, transcriptional, and post-transcriptional levels. Among the many mechanisms that human cells employ to silence repeat expression, maintenance of epigenetic marks on DNA and histones is crucial to healthy brain development.

Dysregulation of repeat-derived RNAs and proteins has been reported in neurological diseases including neurodegenerative diseases such as Alzheimer’s disease, Parkinson’s disease, and multiple sclerosis (MS) (Evans & Erwin, 2021). A series of studies investigating the role of repeat-derived products in MS found that human endogenous retroviruses, also known as HERVs, are increased in several patient samples and are associated with disease status (Macías-Redondo et al., 2021).

Our current understanding of repeat expression and age relies on limited data from non-human model systems, peripheral tissues, and postmortem tissue from limited developmental timepoints (Bogu et al., 2019; de Cecco et al., 2013; He et al., 2021; Lee et al., 2012; Li et al., 2013; Schmitt et al., 2013). As a result, the field has struggled to contextualize changes to repeat expression in neurological disease states without a comprehensive understanding of repeat expression across age.

This same hypothesis has been applied to aging, termed sterile inflammation, and suggests repeat-derived RNAs contribute to a positive feedback loop of functional and biochemical decline with age (Dumetier et al., 2022; López-Otín et al., 2013). Dysregulation of epigenetic machinery is a hallmark of aging, and one theory suggests aging induces a global relaxation of heterochromatin. Thus, remodeling of the epigenetic landscape confers transcriptomic changes and a global de-repression of repeats (LaRocca et al., 2020). The link between repeat expression and aging is evidenced by several observations. Patients with progeroid syndromes, genetic diseases that mimic physiological aging, exhibit increased LINE-1 repeat expression (Della Valle et al., 2022). Furthermore, prematurely aged mice, exhibiting hallmarks of aged chromatin, showed increased LINE-1 (long interspersed nuclear elements) repeat expression (Simon et al., 2019). Upon treatment with reverse transcriptase inhibitors, compounds that target LINE-1 RNA products from being reverse transcribed, mice showed increased health and longevity. Together, these findings suggest repeat derepression contributes to aging phenotypes.

These observations converge in Alzheimer’s Disease, a spontaneous/idiopathic neurodegenerative disease where disease risk is associated with age (Guerreiro & Bras, 2015). Tau neurofibrillary tangles, a neuropathological hallmark of AD, has been implicated as causative in AD though the mechanism is unclear. Studies report that tau promotes neurodegeneration through chromatin relaxation and thus, activates repeat expression in human postmortem brains (Frost et al., 2014; Guo et al., 2018). It is still unclear how repeat expression contributes to disease etiology, but it is possible repeat expression is a feature of aging that is exacerbated by disease.

Our study leverages a large postmortem transcriptome cohort spanning the human lifespan to assess repeat expression in three neurotypical brain regions – caudate nucleus (n=271), dorsolateral prefrontal cortex (DLPFC; n=304), and hippocampus (n=310). We pay particular attention to the expression of repeats associated with human health and disease, specifically LINEs, long terminal repeats (LTRs), short interspersed nuclear elements (SINEs), and satellite repeats. By employing co-expression and differential expression analyses across the three brain regions as a function of age, we glean valuable insights into the region-specific dynamics of repeat expression during aging and its links to age-related neurological disorders.

## Results

### Selection of repeat quantification method

Here, we aimed to quantify repeat-derived RNAs from 885 postmortem brain samples (n=291 female, n=594 male; **Supplemental Figure 1A**). To build this resource of repeat-derived RNA expression across three brain regions (caudate nucleus [n=271], DLPFC [n=304], and hippocampus [n=310]) (Benjamin et al., 2022; Jaffe et al., 2018; Schubert et al., 2015), we first tested two distinct methods designed to quantify repeat expression, TEcount (Jin et al., 2015) and featureCounts (Liao et al., 2014). Both featureCounts and TEcount quantify genomic features (genes and repeats) and share 2,336,733 unique repeat sequences (**Supplemental Figure 1B**).

However, when considering all quantifiable repeats, we observe differences between the methods’ potential benefits as featureCounts can quantify genomic features on the negative strand and when considering all repeats, we observed substantial differences between the two methods with about 50% of the repeats unique to each method (**Supplemental Figure 1C).** TEcount method is uniquely designed to quantify transposable elements, as reflected in the composition of its annotation file (**Supplemental Figure 1D**). For featureCounts, we generated a custom annotation file similar to TEcounts, using RepeatMasker but allowing for strand-specific information and additional satellite repeat annotations (**Supplemental Figure 1E**).

Despite this, the methods show high Pearsons’s correlation, thus perform similarly, when quantifying evolutionarily young repeats such as L1HS (R^2^=0.98) and SVA F (R^2^=0.89) (**Supplemental Figure 1F-G**). Due to its ability to quantify simple and satellite repeats, including those previously associated with Huntington’s disease, amyotrophic lateral sclerosis, and frontotemporal dementia (Kmetzsch et al., 2020), featureCounts was selected for repeat quantification and downstream analyses. The authors welcome questions to help generate a custom annotation file.

### Quantification of repeat expression across brain age

To generate repeat counts with featureCounts from 885 brain samples spanning prenatal – 90 years across the caudate nucleus (n=271), DLPFC (n=304), and hippocampus (n=310) (**Figure 1A, 1B**), we used a custom annotation file (GTF) generated from University of California Santa Cruz’s RepeatMasker containing stranded information about genes and repeats. Using this custom GTF, featureCounts randomly assigns multi-mapping reads from aligned.bam files to a corresponding genomic feature (**Figure 1C**). Subsequently, quality control was performed, and an expression matrix was generated containing 268 caudate samples, 287 DLPFC, and 306 hippocampus samples (**Supplemental Table S1**).

**Figure 1.**
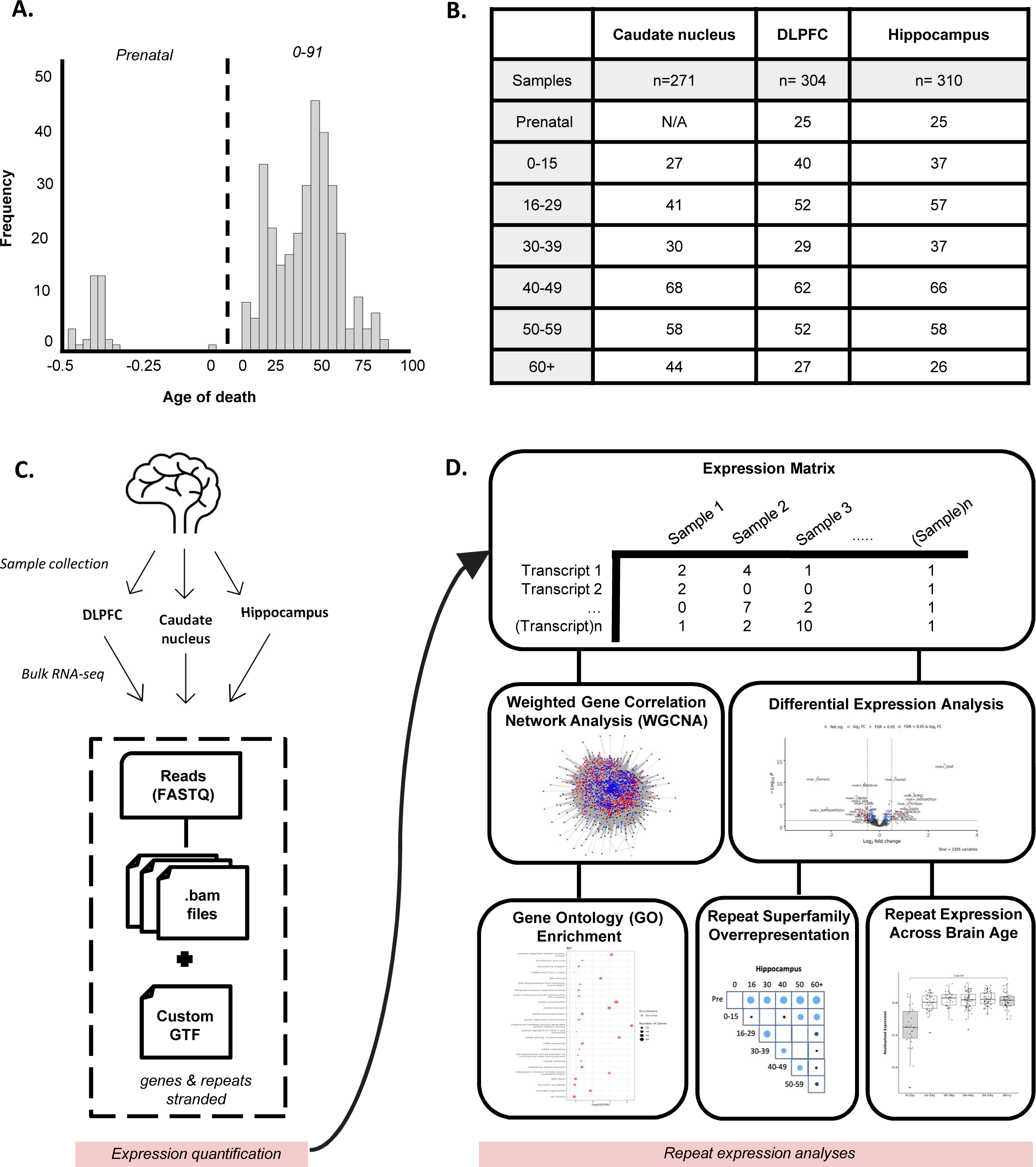
Characterizing repeat expression over lifespan of the neurotypical postmortem brain. A) Neurotypical postmortem brain samples (n=855) included in study span from prenatal to 91 years of age. B) Samples originate from caudate nucleus (n=271), DLPFC (n=304), and hippocampus (n=310) and were binned according to age of death. C) Overview of repeat and gene expression quantification utilizing featureCounts algorithm and custom annotated gene transfer format (GTF) file with hg38 genome. D) Overview of downstream characterization for co-expression analysis (weighted gene correlation network analysis [WGCNA]) and differential expression analysis.

This expression matrix contained quantification of repeat sequences across all samples and was the basis for downstream analyses including differential expression and Weighted Gene Correlation Network Analysis (WGCNA; **Figure 1D**) (Langfelder & Horvath, 2008). Given the high sequence similarity and multiple copies of repeats, along with the ambiguity of short reads, we acknowledge the likelihood for repetitive sequences to map to multiple locations in the genome. Therefore, downstream analyses investigate repeat expression at a superfamily or class level rather than the behavior of individual repeats at specific genomic loci/coordinates.

### Hippocampal repeat expression correlates with brain age

Age-related epigenetic modifications may lead to widespread activation of repetitive elements, with a positive correlation observed between total repeat expression and chronological age. To test this hypothesis, we correlated total repeat expression with ages (0-91 years) across brain regions. Here, we found a slight positive correlation between total repeat expression and age (Figure 2A; R=0.14, p=2.4e-5). Interestingly, we see a minor correlation between total repeat expression and age of death (**Figure 2A**; R=0.14, p=2.4e-5). When we stratify this correlation by brain region, we observe the hippocampus has the highest correlation between total repeat expression and brain age (**Figure 2B**; R=0.17, p=0.0024; Spearman Rank Correlation). The caudate nucleus and DLPFC had negligible correlation. Taken together, this data suggests a relationship between age and repeat expression that is unique to neurotypical hippocampus.

**Figure 2.**
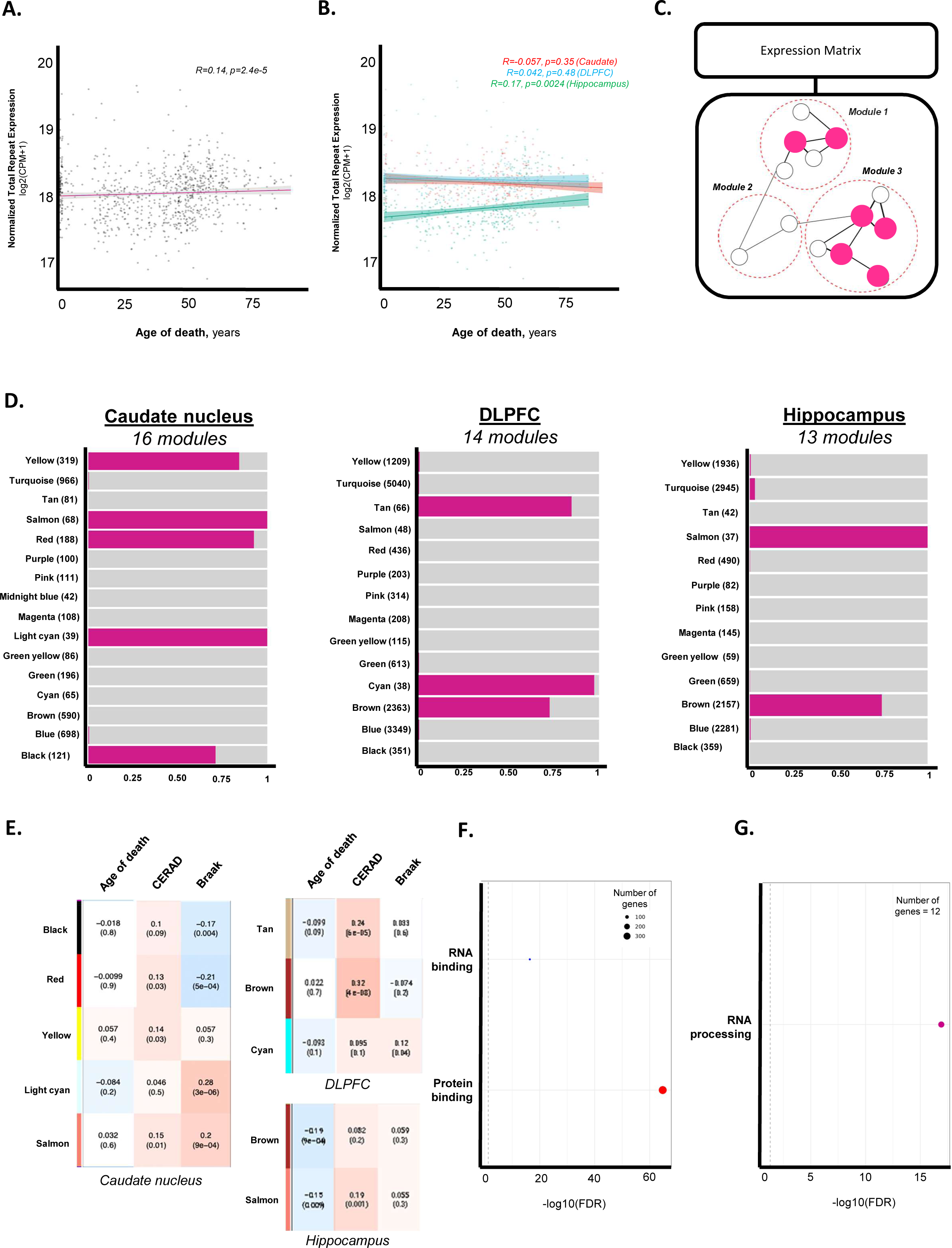
Repeats cluster together by co-expression and genes associated with RNA biology. A) Total repeat expression from uniquely mapped reads mildly correlates with age of death (Spearman two-sided; rho=0.14, p=2.4e-5) B) When stratified by brain region, correlation of total repeat expression with age of is driven by the hippocampus (Spearman two-sided; rho=0.17, p=0.0024) C) Schematic representing how WGCNA utilizes expression matrix to place repeats and genes into neighborhoods and generate co-expression networks. D) WGCNA module composition across caudate nucleus, DLPFC, and hippocampus as a proportion of genes and repeats. E) Heatmap of correlation between repeat-dense modules and clinical traits of caudate nucleus, DLPFC, and hippocampus samples. Repeat-dense modules negatively correlate with age of death across repeat-dense modules. Yet, repeat-dense modules positively correlate with neuropathological scores in the caudate nucleus. F) Gene ontology enrichment for DLPFC Brown module. G) Gene ontology enrichment for caudate nucleus, Light Cyan module.

### Repeats co-expression with RNA binding genes

To understand the biological pathways associated with repeats and age across the neurotypical brain, we performed weighted gene co-expression network analysis (WGCNA) on both gene and repeat expression profiles derived from bulk RNA-seq data for each of the three brain regions. WGCNA utilizes the gene expression matrix generated from uniquely mapped reads to cluster expressed features (repeats or genes) into co-expression modules, referred to as modules in the remainder of the text, based on the Pearson correlation coefficient between a pair of features. Previously, this method has been applied to gene expression, miRNA expression and DNA methylation (Euclydes et al., 2022; Langfelder & Horvath, 2008; Pascut et al., 2020), but here, we apply WGCNA to a residualized expression matrix containing both repeat and gene expression. To reduce artifacts introduced by evolutionarily young repeats, only uniquely mapped reads were considered (**Methods**) when generating brain region networks using power **β = 14** threshold (**Supplemental Table 2**). Given the potentially region-specific relationship between repeat expression and age, we applied WGCNA to organize 2336 unique repeats and 21986 unique genes into modules by brain region to generate three regional co-expression networks (**Figure 2C**). The caudate nucleus, DLPFC, and hippocampus produced 16, 14, 13 modules, respectively. Module size varies and the total number of features (repeats and genes) within a module is indicated in parentheses and is detailed in (**Supplemental Table 3**).

To investigate the heterogeneity of each module, we visualized the proportion of repeats and genes. Modules composed of >50% of total features were classified as repeat-dense. The number of repeat-dense modules varied by brain region (**Figure 2D; Supplemental Table 3**). Interestingly, while it had the highest correlation between total repeat expression and age, the hippocampus had the fewest number (2) of repeat-dense modules compared to caudate nucleus (5) or DLPFC (3).

To evaluate if repeat-dense clusters correlate with age and others clinical features related with senescence, we calculated the Pearson correlation coefficient between each WGCNA’s module eigengene (kME) and our clinical features. Sample characteristics selected include age, as well as neuropathological neuritic plaque (CERAD) and neurofibrillary tangle (Braak) scores (**Supplemental Table 1**). Both CERAD and Braak scales are neuropathological metrics of neurodegeneration associated with Alzheimer’s and Parkinson’s Disease, respectively (Burke et al., 2008; Fillenbaum et al., 2008). Repeat-dense modules had mild correlations of <|0.2| with age of death, with DLPFC’s Brown and Salmon modules having p-value=9e-04 and p-value=0.009, respectively (**Figure 2E**). Both hippocampus’ repeat-dense modules had a negative correlation with age of death at (R=-0.19, p=9e-04; Pearson correlation) and (R-0.15, p=0.009; Pearson correlation), respectively. In contrast, repeat-dense modules positively correlate (>0.2) with both CERAD and Braak scores in both the caudate nucleus and DLPFC (**Figure 2E**). In the caudate, the Light Cyan module correlates moderately with Braak score (R=0.28, p=3e-6; Pearson correlation) and is composed of 100% repeats. In the DLPFC, the Brown module correlates moderately with CERAD score (R=0.32, p=4e-8; Pearson correlation).

The DLPFC Brown module is composed of 72.71% repeats and 27.29% genes. We next assessed the biological function of the genes contained in these repeat-dense modules. Thus, we performed gene ontology (GO) enrichment analysis on all clusters to identify enrichment of molecular function, biological processes, and cellular compartments with particular interest in GO enrichment of repeat-dense clusters containing genes (**Supplemental Table 4**). GO analysis of the DLPFC Brown module, caudate Light Cyan module, and caudate Yellow module revealed enrichment of genes associated with RNA and protein binding, RNA processing, and molecular functions associated with neurodegeneration including ubiquitin-protein transferase activity and ubiquitin conjugating enzyme (**Figure 2E, 2F; Supplemental Figure 3A**). These results suggest repeats correlate with clinical metrics of neurodegeneration.

### Age-associated gene modules enriched for immune response and transcriptional regulation

Gene expression is known to change with age (de Magalhães et al., 2009), therefore, we also investigated GO enrichment of gene-dense modules that correlate with age of death. There were several gene-dense clusters that have a strong, positively correlated relationship with age of death including DLPFC’s blue and Hippocampus’ yellow module **(Supplemental Table 3, 4**). The DLPFC’s Blue module (n= 3329 genes and 20 repeats) correlated negatively with age of death (R=-0.40, p=2e-12) was enriched for genes associated with transcriptional regulation by RNA Pol II (**Supplemental Figure 3B**). The hippocampus’ Yellow module (n=1925 genes and 11 repeats) correlated positively with age of death (R=0.55, p=2e-25) and was enriched for genes associated with immune response, inflammatory response, and defense response to virus (**Supplemental Figure 3C**). Together, these results support that gene-dense modules hold meaningful information about brain age.

### Identification of differentially expressed repeats across lifespan of neurotypical brain: brain regions show distinct repeat family association with aging

We hypothesized a differential expression (DE) analysis, comparing repeat expression between over age, could identify differentially expressed repeats (DER) that are potential biomarkers of the neurotypical aging brain. To perform this analysis, we utilized samples binned by age (Prenatal; 0-15; 16-29; 30-39; 40-49; 50-59; 60+) where each age bin contained an adequate sample size with a minimum of 25 samples. (**Figure 1B, Supplemental Table 5**). Prenatal samples were only available for hippocampus and DLPFC.

Using counts generated with featureCounts (**Methods**), residualized expression was generated using covariates via voom linear model. In this analysis, we controlled for the effect of biological sex, self-reported race, ancestry (SNP PCs 1-3), and RNA quality (RIN, mitochondria mapping rate, gene assignment rate, genome mapping rate, and hidden effects using surrogate variable analysis) (**Methods; Model 1, Differential Expression Analyses, Eq.5**), with age as our variable of interest. Criteria for a differentially expressed repeat (DER) was a false discovery rate (FDR) < 0.05. Differential expression identified 21696 DERs across all three brain regions representing 16.58% of all repeats analyzed (**Supplemental Figure 4A, Supplemental Table 6**). For contrast, differential expression analysis from TEcount resulted in 13489 DERs (**Supplemental Table 7**).

Given age is a primary risk factor for many neurodegenerative diseases, and prenatal samples were not available for caudate nucleus, we further investigated DERs between 0-15 vs. 60+ years of age, identifying 1,401 instances between ages 0-15 and 60+ across all three brain regions (**Figure 3A**). When stratifying DERs by significance (FDR<0.05) and magnitude of change, we observed the majority of young (0-15 years) versus older (60+ years) DERs have a relatively small fold change (log2(fold change) < 0.5) across all three brain regions (**Figure 3B**).

**Figure 3.**
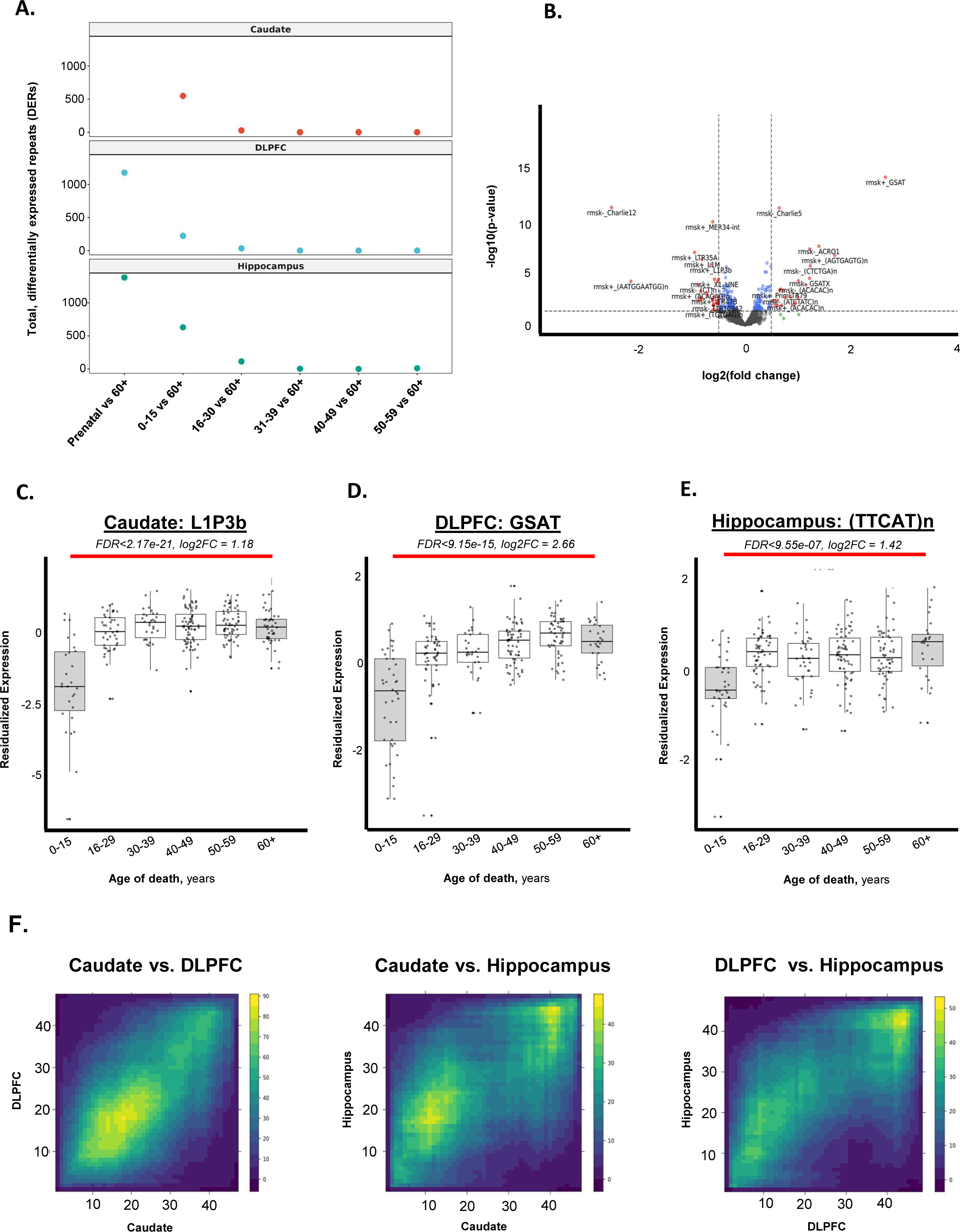
Repeats are differentially expressed between human brains 0-15 vs. 60+ years. A) Total number of DERs across differential expression comparisons with brains >60 years. B) Volcano plot visualizing 0-15 vs. 60y+ DERs by fold change and FDR. C) Most significant 0-15 vs. 60+ DER in caudate nucleus, L1P3b (FDR=2.17e-21, log2(fold change)=1.18). D) Most significant 0-15 vs. 60+ DER in DLPFC, GSAT (FDR=9.15e-15, log2(fold change)=2.66). E) Most significant 0-15 vs. 60+ DER in hippocampus, TTCATn (FDR=9.55e-7, log2(fold change)=1.42). F) R-RHO plot depicting concordance between 0-15 vs. 60y+ DERs between brain regions.

Across the caudate nucleus, DLPFC, and hippocampus, the most significant DERs between young (0-15 years) and older (60+ years) individuals belonged to distinct repeat families. In the caudate nucleus, the most upregulated DER was the LINE repeat L1P3b (FDR=2.17e-21). The DLPFC showed the greatest increase in centromeric GSAT repeats (FDR=9.15e-15), while the hippocampus exhibited the strongest upregulation of the satellite repeat TTCATn (FDR=9.55e-7) (Figure 3C-E). To explore consistency in DER patterns across brain regions, we employed Rank-Rank Hypergeometric Overlap (RRHO) analysis (Plaisier et al., 2010). This analysis revealed the strongest concordance in the direction of change (up or downregulation) between DERs in the caudate nucleus and DLPFC (Figure 3F). Concordance was weaker between the DLPFC and hippocampus, and the hippocampus and caudate nucleus. All three brain regions exhibited an increase in expression from young (0-15 years) to older (60+ years) groups (**Supplemental Figure 4B**).

### Age-related repeat expression is brain region specific

Discussions of repeat expression are often centered around increases in repeat expression, as it poses threat to genomic stability and cellular homeostasis. In our analysis, we unbiasedly captured DERs with FDR<0.05, regardless of directionality of change. To test whether repeat expression ubiquitously increases with age, we stratified 0-15 vs. 60+ DERs by the sign (positive or negative) of log2(fold change) (**Supplemental Table 6**). We identified 549 DERs (403 upregulated, 146 downregulated) in caudate nucleus, 233 DERs (92 upregulated, 233 downregulated) in DLPFC, and 629 DERs (25 upregulated, 629 downregulated) in hippocampus (**Supplemental Figure 5A, 5B**). While 73.40% of caudate nucleus DERs are upregulated in brains >60 years, 96.02% hippocampus DERs are downregulated in brains >60 years.

To test if the observations were representative across multiple categories of repeats, we investigated the behavior of LINEs, LTRs, SINEs, and satellite repeats present within 0-15 v. 60+ DERs (**Figure 4A-D**). LINEs and LTRs were the most abundant categories present within 0-15 vs. 60y+ DERs and shared similar patterns of upregulation in caudate nucleus >60y and downregulation in hippocampus >60 years (**Figure 4A, 4B; Supplemental Table S6**). Interestingly, despite moderate correlation between total repeat expression and age in the hippocampus (R=0.17; **Figure 2B**), hippocampus DERs are primarily downregulated in brains between 0-15 and 60+ years. This suggests a small subset of non-differentially expressed repeats may drive global correlation of repeat expression-age and are not representative of more nuanced changes in repeat expression with age.

**Figure 4.**
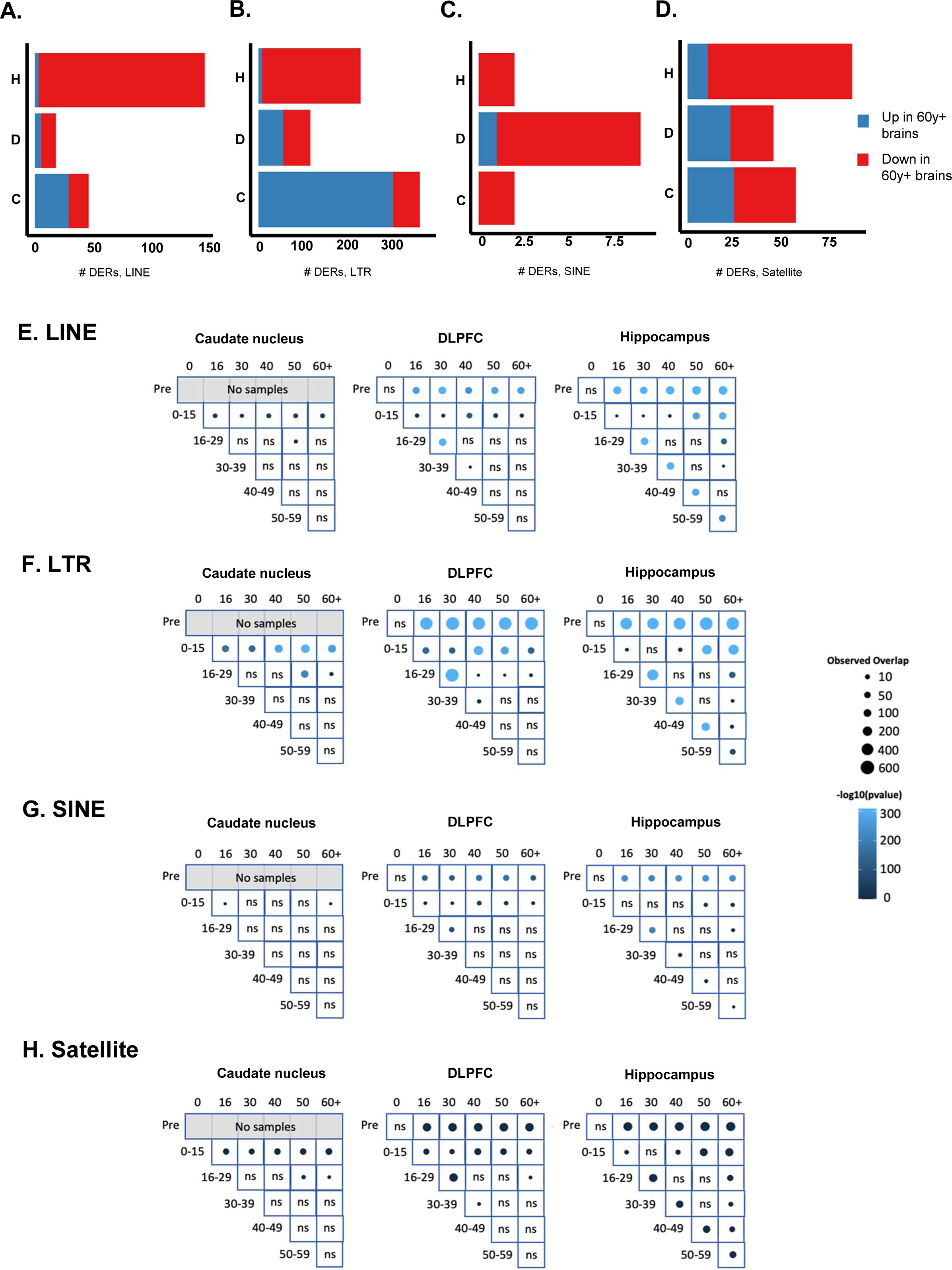
LTRs are over-represented in differential expression results. A-D) Total number of 0-15 vs. 60+ DERs downregulated (red) vs upregulated (blue), stratified by LINE, SINE, LTR, and satellite repeats E-H) Over representation analysis of LINE, SINE, LTR, and satellite repeats within DER results across all comparisons and brain regions (DER = differentially expressed repeat, LINE = Long Interspersed Element, LTR = Long Terminal Repeat, SINE=Short Interspersed Repeat)

### LTRs are over-represented within differential expression results

Given the unique behavior of differentially expressed repeats across brain regions, we wanted to see if any repeat classes were more likely to be differentially expressed. To test if LINEs, LTRs, SINEs, or satellite repeats were over-represented within our differential expression results, we performed the Hypergeometric distribution test, implemented by the SuperExactTest package. This methodology calculates the significance of the intersection between two sets of elements (repeats), considering a fixed background population (all unique annotated repeats, **Methods**). With this approach, we can assess if a specific differentially expressed repeat is overrepresented within its own repeat super family, compared to what would be expected by chance, considering all repeats annotated in our custom.gtf file (**Methods**). Results from each individual comparison performed across all three brain regions are shown in **Figure 4E-H**. We quickly identified a striking over-representation of LTR elements within our differential expression results including within 0-15 vs. 60y comparison across all three brain regions (**Figure 4F**).

### Age contributes to differential expression of HERV-K in brain

Given LTRs are abundant and overrepresented within 0-15 vs 60+ DERs across all three brain regions, we then wanted to evaluate if any differentially expressed LTRs were shared between the caudate nucleus, DLPFC, and hippocampus. Out of the 1401 0-15 vs 60+ DERs identified, 36 DERs were shared between all three tissues (**Figure 5A**). Of these, 17 were LTRs and 10 were satellite repeats (**Figure 5B**). Further investigation into 17 shared LTRs yielded identification of human endogenous retrovirus-K-int (HERV-K-internal sequences), a human-specific LTR, as a significant 0-15 vs 60y+ DER across caudate nucleus (FDR=0.011;), DLPFC (FDR=0.043), and hippocampus (FDR=0.00026) (**Figure 5C**).

**Figure 5.**
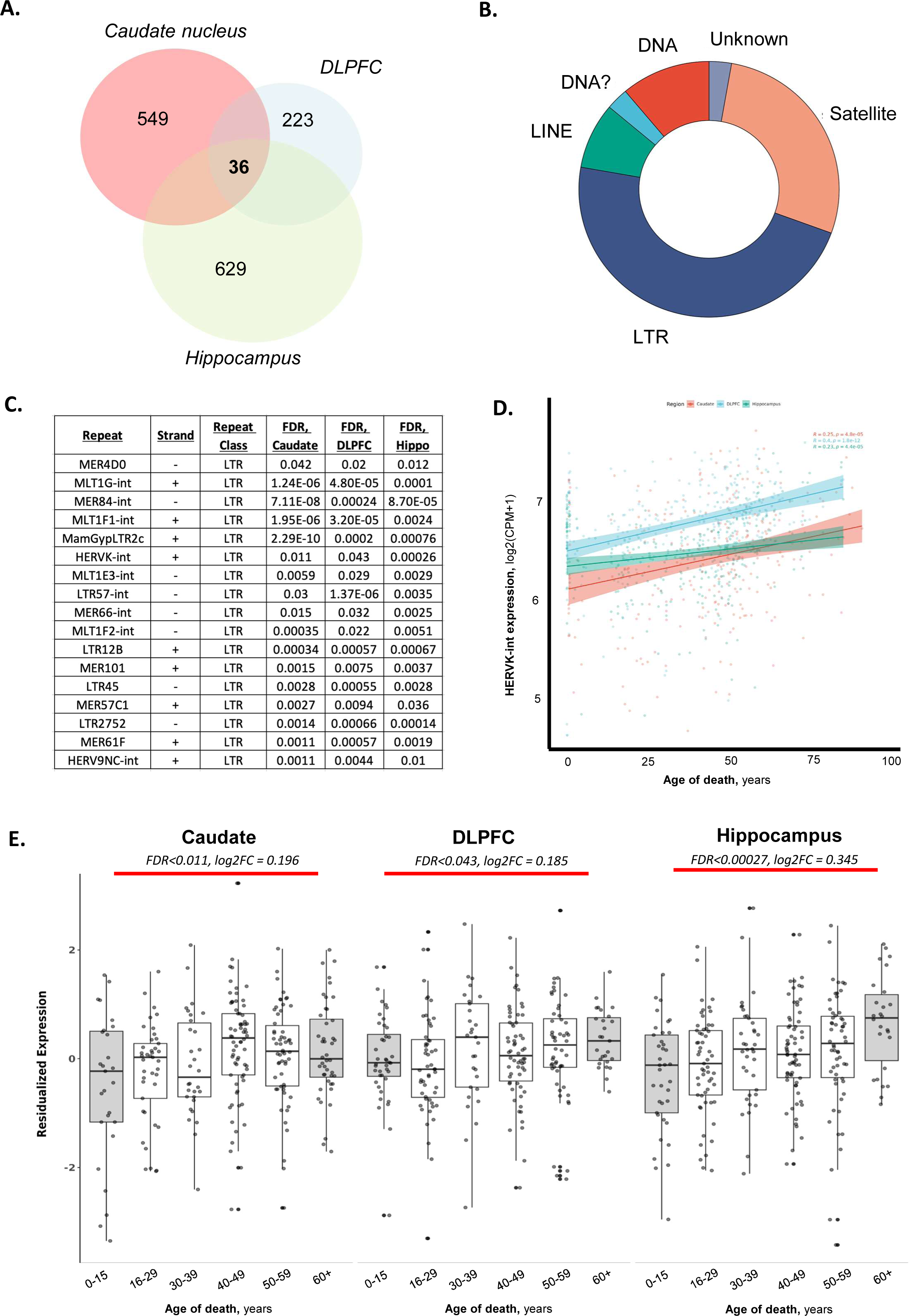
HERV-K expression increases from 0-15 vs. 60+ years in all three brain regions. A) Venn diagram of 0-15 vs. 60+ DERs across brain regions B) Distribution of repeat classes within shared 0-15 vs. 60+ DERs across all three tissues C) List of 17 LTR 0-15 vs. 60+ DERs shared across all three tissues D) Correlation between total expression of HERV-K-int and age of death by brain region E) Expression of HERV-K-int across age bins, by brain region.

Previously, HERVs have been identified as being dysregulated in neurological disorders including multiple sclerosis (Brudek et al., 2009; Laufer et al., 2009; Schmitt et al., 2013). Age is a risk factor for multiple sclerosis, thus, we asked if total LTR, HERV, or HERV-K expression correlated with brain age in our samples. We observed that total LTR expression has the highest Spearman Rank Correlation (R=0.077, p=0.024; test) out of all repeat categories analyzed (**Supplemental Figure 6A**). When we look at total HERV-K expression, a subcategory of LTR elements, we see HERV only mildly expression correlates with brain age (R=0.033, p=0.0024; test) and when plotted by brain region, is similar across caudate nucleus (R=0.068, p=0.00045), DLPFC (R=0.051, p=0.0059), and hippocampus (R=0.044, p=0.014) **Supplemental Figure 6B, 6C**). Importantly, when plotting total HERV-K expression, we observe a strong and significant positive correlation with brain age across all three brain regions. Total expression of HERV-K has the highest correlation with brain age in the DLPFC (**Figure 5D**, R=0.4, p=1.8e-12). Boxplots further confirm HERV-K expression increases with age in the neurotypical caudate nucleus, DLPFC, and hippocampus (**Figure 5E**).

Re-investigating the co-expression analysis, we observe HERV-K-int clusters into the DLPFC Brown module containing 1718 repeats and 645 genes (**Supplemental Table XX**). Of the 1718 repeats, 61 (3.55%) are both human endogenous retrovirus and a highly connected feature (hub) of the DLPFC Brown network. Of the 61 repeats that serve as hubs, 13 (21.3%) are from HERV-K subfamily, including HERV-K-int. All HERV-K hubs are all classified as intramodular hubs, hubs that are highly connected within the DLPFC Brown module and drive architecture of network, as indicated by a high kWithin and positive kDiff values (**Supplemental Table S8,** Bogenpohl et al., 2016). As previously mentioned, the DLPFC Brown module shows a significant positive correlation with CERAD score (r=0.32, p<4e-08) (**Figure 2E**) indicating a potential relationship with HERV-K expression and neuropathological protein aggregation.

GO analysis of genes in the DLPFC Brown module are enriched for molecular functions of protein and RNA binding (**Figure 2F**). Together, this data suggests expression of HERV-K-int correlates with brain age and shares co-expression patterns with several pathways critical to cellular homeostasis that have been previously implicated in the aging brain (Ham & Lee, 2020).

## Discussion

In this study, we re-processed and re-quantified paired-end, stranded RNA-sequencing data from 885 neurotypical samples across the caudate nucleus, DLPFC, and hippocampus from 395 human, postmortem BrainSeq consortium donors (Benjamin et al., 2022; Collado-Torres et al., 2019) to build a repeat expression atlas of the aging human brain. Using co-expression networks, we placed repeat-derived RNAs within the brain’s transcriptional network to discover biologically relevant relationships between repeats and genes.

The impact of genomic instability, epigenetic alterations, and altered cellular communication, all hallmarks of aging, are not exclusive to genes and likely impact global expression (López-Otín et al., 2013; Yamamoto et al., 2022). The interconnectedness between repeat and gene expression has largely been understudied in the context of aging until recently. Through WGCNA, we identified that repeats generally cluster together into repeat-dense modules, an expected result given that repeats are the target of shared regulatory mechanisms at the transcriptional and post-transcriptional levels. Gene-dense modules appear to hold more information about brain age, an expected result given previous studies on age-related expression changes in the brain. Interestingly, repeat-dense modules correlate with CERAD and Braak scores, suggesting a relationship between repeat expression and neuropathological hallmarks of disease, independent of age (Yamamoto et al., 2022). For example, the repeats in Light Cyan module in caudate consists exclusively of LINE-1 elements, sequences harboring conserved potential G-quadruplex (G4) forming sequences in their 3’ end which are associated with increased retrotransposition (Sahakyan et al., 2017). Notably, LINE-1s have been associated with G4 formation in Alzheimer’s disease induced pluripotent stem cells derived neurons (Hanna et al., 2021), being an intragenic feature reducing gene expression and potentially affecting the transcriptional programs.

We then went on to identify 21696 DERs (FDR<0.05) across the caudate nucleus, DLPFC, and hippocampus with the DLPFC containing the fewest DERs, mimicking differential expression results reported in Collado-Torres et al., 2019. We observed an overrepresentation of LTRs within our DER results and among our 0-15 vs. 60y+ DER results, we identified 17 LTRs that were shared across caudate nucleus, DLPFC, and hippocampus. Of these 17, we observed HERV-K increases with brain age and is upregulated in brains >60 years.

While we do not observe a strong global relationship between repeat expression and age, we identified HERV-K as a repeat signature associated with the aging neurotypical brain. Not only is HERV-K-int a shared DER across all three brain regions, but its total expression is moderately correlated with brain age and stronger than the correlation of total expression HERV-K element expression with brain age. A recent study supports the connection between endogenous retrovirus expression and cellular senescence indicating HERV-derived proteins, including HERV-K, can serve as a biomarker of tissue aging across lung, liver, and skin (36610399). Our study expands upon this observation to confirm HERV-K RNA is a biomarker of aging across the brain, broadening and strengthening HERV-K’s position in the diagnostic and therapeutic landscape of age-related neurodegeneration.

LTRs, more specifically HERVs, have also been associated with neurological disease (Dembny et al., 2020; Macías-Redondo et al., 2021). HERV-derived RNA is capable of causing and propagating neurodegeneration through Toll-like receptors (Dembny et al., 2020) and protein aggregation (Liu et al., 2023). Beyond the production HERV RNA species, Turelli et al. observed the regulatory impact of HERV-K also stems harboring transposable element-embedded regulatory sequences (TEeRS) and subsequently altering KRAB-ZFP, a transcriptional repressor, binding to neuronal genes (Turelli et al. 2020). We observe HERV-K-int, along with other HERV-K sequences, are intramodular hubs within our DLPFC and hippocampus co-expression networks suggesting this LTR maintain and/or regulate biologically important relationships within a brain region. Repeat-derived products, including repeat-derived RNAs, are not only capable of propagating neurodegeneration through a pro-inflammatory response – thus contributing to disease progression (Dembny et al., 2020); but also in some forms of cancer through the ubiquitin-proteasome pathway (Jin et al., 2019) and on aging phenotype (Gorbunova et al., 2021). Thus, we also propose elevated HERV-K products in neurological disease may reflect a molecular phenotype of accelerated aging that further drives transcriptional and proteomic hallmarks of neurodegeneration.

Comprehensively, our work provides the largest global assessment of repeat expression across the aging neurotypical brain and refutes previous generalizations of repeat behavior. While epigenetic alterations may change transcriptional landscape with age, we find repeat expression shows high developmental and regional specificity making age only one important factor for characterizing repeat behavior in a healthy aging tissue. We hope this global assessment will serve as a resource to the greater scientific community.

Thus, as repeat expression becomes a popular target for biomarkers, diagnostics, and therapeutics, our findings highlight the need to identify baseline expression dynamics of target repeats in healthy tissues. As such, we anticipate this data will be used as a neurotypical baseline for analyzing neurodevelopmental and neurodegenerative disease-related changes in repeat expression.

## Methods

### Sample Selection

The LIBD BrainSeq Consortium consists of several brain regions and includes a wide range of demographics and RNA-sequencing library preparation. We selected samples from the caudate nucleus, DLPFC, and hippocampus based on three inclusion criteria: 1) Stranded RiboZero RNA-sequencing library preparation, 2) primary diagnosis of neurotypical control, and 3) self-reported ancestry of either African American or European American. This resulted in a total of 395 unique individuals for a total of 885 FASTQ files across the three brain regions.

### RNA-sequencing Data Processing

We downloaded FASTQ files from the BrainSeq Consortium (Benjamin et al., 2022; Jaffe et al., 2018; Schubert et al., 2015). The reads were aligned to the hg38/GRCh38 human genome (GENCODE release 26, GRCh38.p10) using HISAT2 (v2.1.0) (Kim et al., 2019). Following genome alignment, we sorted and indexed the BAM files using SAMtools (v1.9) (Danecek et al., 2021) with HTSlib (v1.9) (Bonfield et al., 2021). We examined alignment and read quality with RSeQC (v3.0.1) (L. Wang et al., 2012). We generated gene and repeat counts from both multi mapping (expression and downstream analysis) and unique mapping (co-expression analysis) using TEtranscripts (v2.2.1) (Jin et al., 2015) and featureCounts (v2.0.1) (Liao et al., 2014).

### Generating Counts from Multi-Mapping Reads and Repeat Annotation

For TEtranscripts, we generated gene and repeat counts in one step using TEcount for paired end, reversed stranded reads with default parameters and the GTF file of genes and repeats provided by the Hammel Lab (http://hammelllab.labsites.cshl.edu/software/). Additionally, we used featureCounts to generate gene and repeat counts in one step with a customized GTF file of genes and repeats. For the GTF file generation, we combined the GENCODE release 26 with repeat annotation obtained from downloading the repeat masker track for hg38 from the UCSC Table Browser followed by annotating strand information with a python script. We generated counts with featureCounts using the following parameters: 1) paired end, 2) reversed stranded reads, 3) primary alignments only, 4) excluding chimeric reads, 5) allowing for multi-mapping reads and, 6) one base as the minimum overlapping fraction in a read.

### Quality Control

For quality control, we first aggregate results for RSeQC and HISAT2 with MultiQC (Ewels et al., 2016). To determine outliers, we first combined the read, alignment, and RNA quality (RIN: RNA Integrity Number and mitochondria mapping rate) for each tissue and scaled the data before applied dimensional reduction with PCA (principal component analysis) with the scikit-learn package (Pedregosa et al., 2011). Following dimensional reduction, we calculated the distance from the centroid for all samples (**Equations 1 & 2**) and excluded samples that were outside of the 99 percentile (caudate and hippocampus) and 95 percentile (DLPFC). This resulted in a total of 861 samples for caudate (n=268), DLPFC (n=287), and hippocampus (n=306).

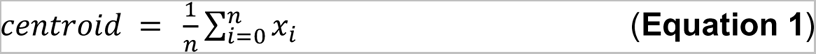

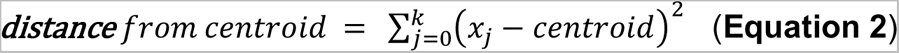

### Low expression filtering and library normalization

To filter out low expression counts, we first constructed an edgeR object (McCarthy et al., 2012; Robinson et al., 2009) of brain regions with sample information as well as raw counts. Following this, we applied filterByExpr (Chen et al., 2016) from edgeR for genes and repeats together with an interacting design matrix (**Equation 3**). This function keeps features (genes and repeats) that have count-per-million (CPM) above a minimum count (10 CPM) in 70% of the smallest group sample size. The smallest group sample size is determined by the design matrix. Furthermore, each feature must have a minimum number of counts across all samples (15 CPM). After filtering, we had a total of 28443, 28058, and 28740 genes and repeats for the caudate nucleus, DLPFC, and hippocampus, respectively for the TEtranscripts method. For the featureCounts methods, this resulted in 24861, 24545, and 25022 genes and repeats for the caudate nucleus, DLPFC, and hippocampus, respectively. After filtering, we normalized the library size of genes and repeats together using trimmed mean of M-values (TMM).

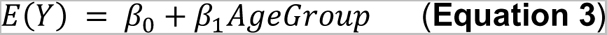

### Repeat Expression Analyses

#### Expression residualization

For residualized expression, we regressed out covariates using limma-voom normalized expression and null models created without the variable of interest (**Equation 5**) as previously described in Benjamin et al. 2022. Following this, a z-score transformation was performed.

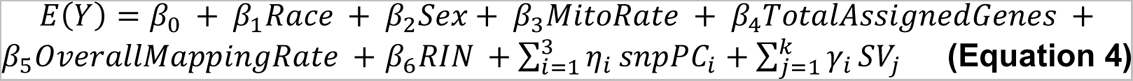

#### Differential expression analyses

For differential expression analyses, we applied voom normalization law (Law et al., 2014) on the normalized filtered counts for genes and repeats together (**Low expression filtering and library normalization**), adjusted with the model covariates listed below (Leek et al., 2012) (**Equation 4**). Differentially expressed features were identified using the eBayes (Smyth; Hall, 2009) function from limma (Ritchie et al., 2015) for the age group fitted model. Features with a FDR < 0.05 were considered as differentially expressed.

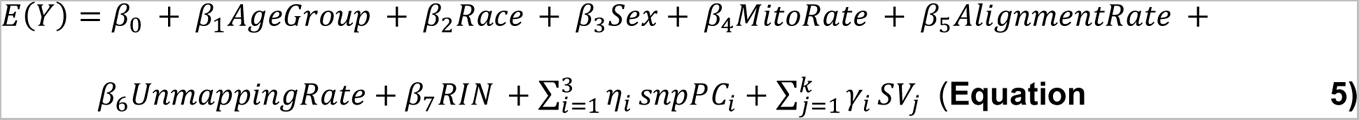

Covariates included sex, self-reported race, ancestry (SNP PCs 1-3), and RNA quality (RIN, mitochondria mapping rate, alignment rate, genome unmapping rate, and hidden variance using surrogate variable analysis (SVA)).

#### Repeat Superfamily Hypergeometric Analysis

To evaluate the representation of repeat superfamilies in each differential expression result, we applied the supertest function from SuperExactTest (Wang et al., 2015). For superfamily analysis, we utilized the total number of repeats (n = 30938) included in the custom GTF file as background population. We then used the intersection between the set of repeats of a repeat superfamily (i.e. all 342 LINEs repeats present in our GTF file) with the total number of differentially expressed repeats within the same repeat family, obtained in each age comparison analysis. We performed the Hypergeometric test in 16 repeat super families. The P-values were calculated by the same function considering only the upper tail of the distribution of each intersection.

### WGCNA analyses

#### Generating Counts from Uniquely Mapped Reads

Considering the uneven distribution of repeats across the genome and high sequence similarity, we also generated counts from uniquely mapped reads to reduce artifacts introduced by evolutionarily young repeats (Parsana et al., 2019). We generated counts with featureCounts using the following parameters: 1) paired end, 2) reversed stranded reads, 3) primary alignments only, 4) excluding chimeric reads, 5) excluding multi-mapping reads and, 6) one base as the minimum overlapping fraction in a read.

#### Co-expression Analysis

We used the Weighted Correlation Network Analysis (WGCNA) to create co-expression networks for each brain region and identify co-expression modules (Langfelder & Horvath, 2008). As an input we used the residualized expression previously described (**Equation 5)**, with genes and repeats together, from featureCounts. We analyzed each brain region in separate, using all age groups from each tissue. We select a β = 14, (a value with all networks achieved a scale –free independence index of R^2^ ≥ 0.8), using the following parameters: signed network, mergecutheight = 0.25, deepsplit = 2, and minimum module size = 30 (Feltrin et al., 2019). Each individual module eigengene value (kME) were correlated with the following co-variables: self-reported race, sex, age of death, RIN, pH, PMI, MitoRate, AlignmentRate, CERAD/BRAAK scores and each one of the 6 age groups. Modules with a Pearson correlation coefficient p-value < 0.05 were considered as significant associations. For the identification of hub genes of each module, we selected all the genes/repeats selected by the WGCNA function intramodularConnectivity().

#### CERAD/BRAAK Scores

A subset of 108 samples representing 5*7* unique individuals were analyzed by a Lieber Institute for Brain Development neuropathologist. All 108 samples were given both a CERAD scores (1-4) and BRAAK score (1-4).

#### Gene term enrichment analysis

For gene term enrichment analysis, we utilized GOATOOLS, a Python package using hypergeometric tests with the Gene Ontology (GO) database (Klopfenstein et al., 2018). The GO database included molecular functions (MF), cellular components (CC), and biological processes (BP), however, MF and BP were primarily used for analyses.

## Code and data availability

Code will be available at https://github.com/orgs/paquolalab.

## Supplemental Figures

**Figure S1.**
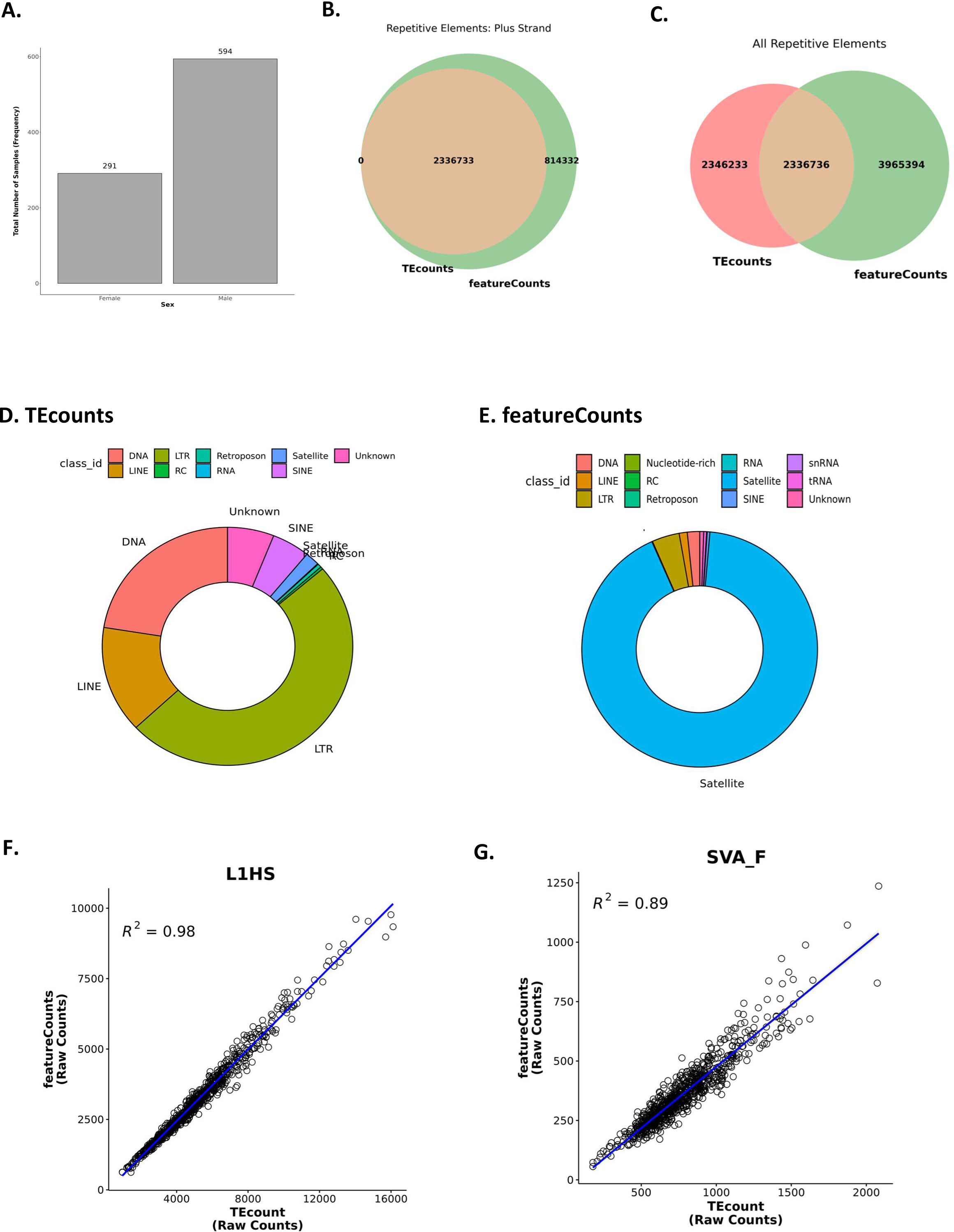
Selection of repeat quantification method. **A**) Breakdown of post-mortem samples by sex. **B)** Overlap of quantifiable repeats on positive strand by featureCounts and TEcounts. **C)** Overlap of all quantifiable repeats features by featureCounts and TEcounts. **D)** Breakdown of categories of quantifiable repeats in TEcounts GTF file. **E)** Breakdown of categories of quantifiable repeats in featureCounts GTF file. **F)** Correlation between featureCounts and TEcounts, raw counts of L1HS. **G)** Correlation between featureCounts and TEcounts, raw counts of SVA_F.

**Figure S2.**
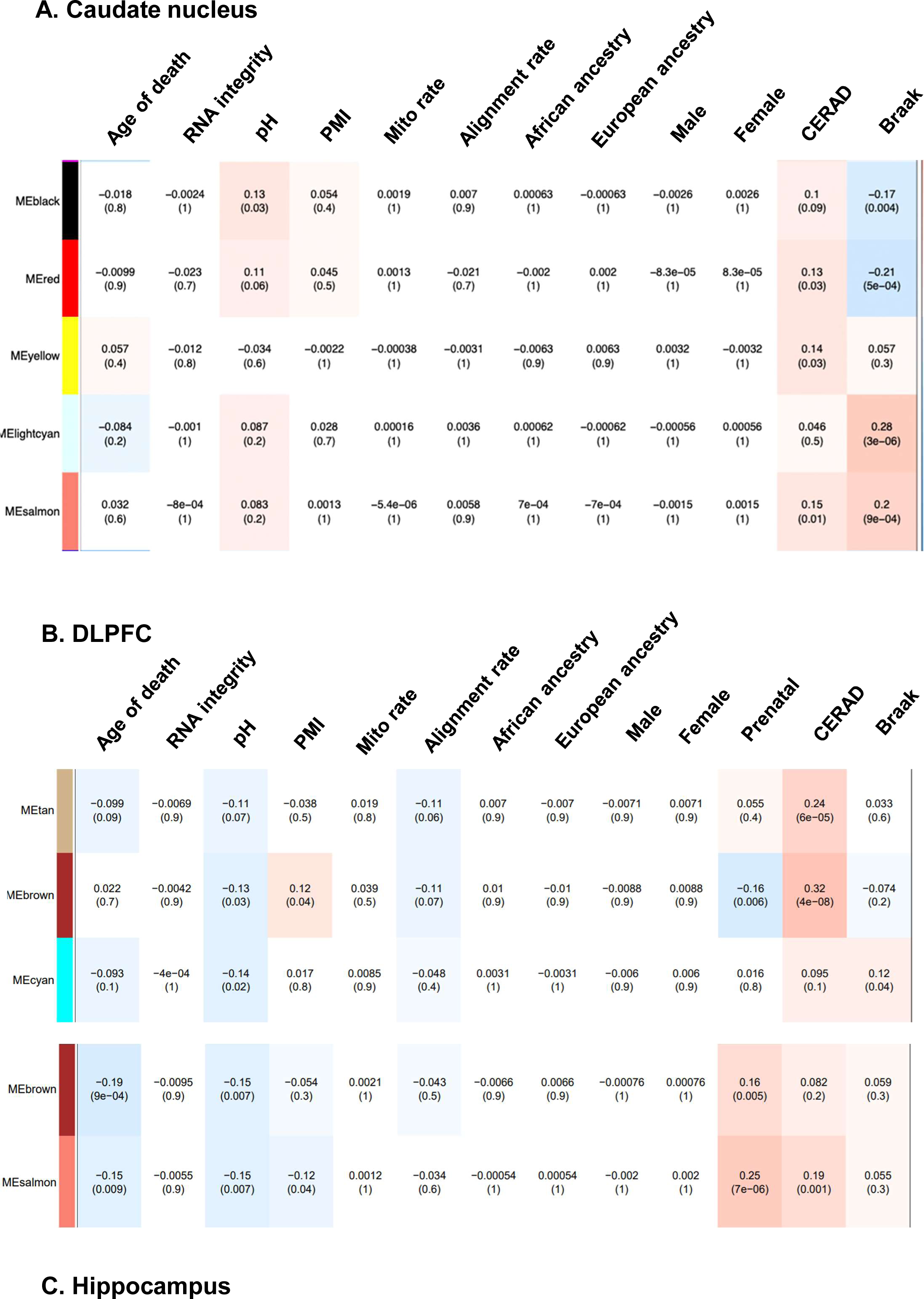
Co-expression defines correlation of repeat-dense WGCNA modules with clinical traits. A) WGCNA module-trait correlation heatmap across caudate nucleus samples. B) WGCNA module-trait correlation heatmap across DLPFC samples. C) WGCNA module-trait correlation heatmap across hippocampus samples.

**Figure S3.**
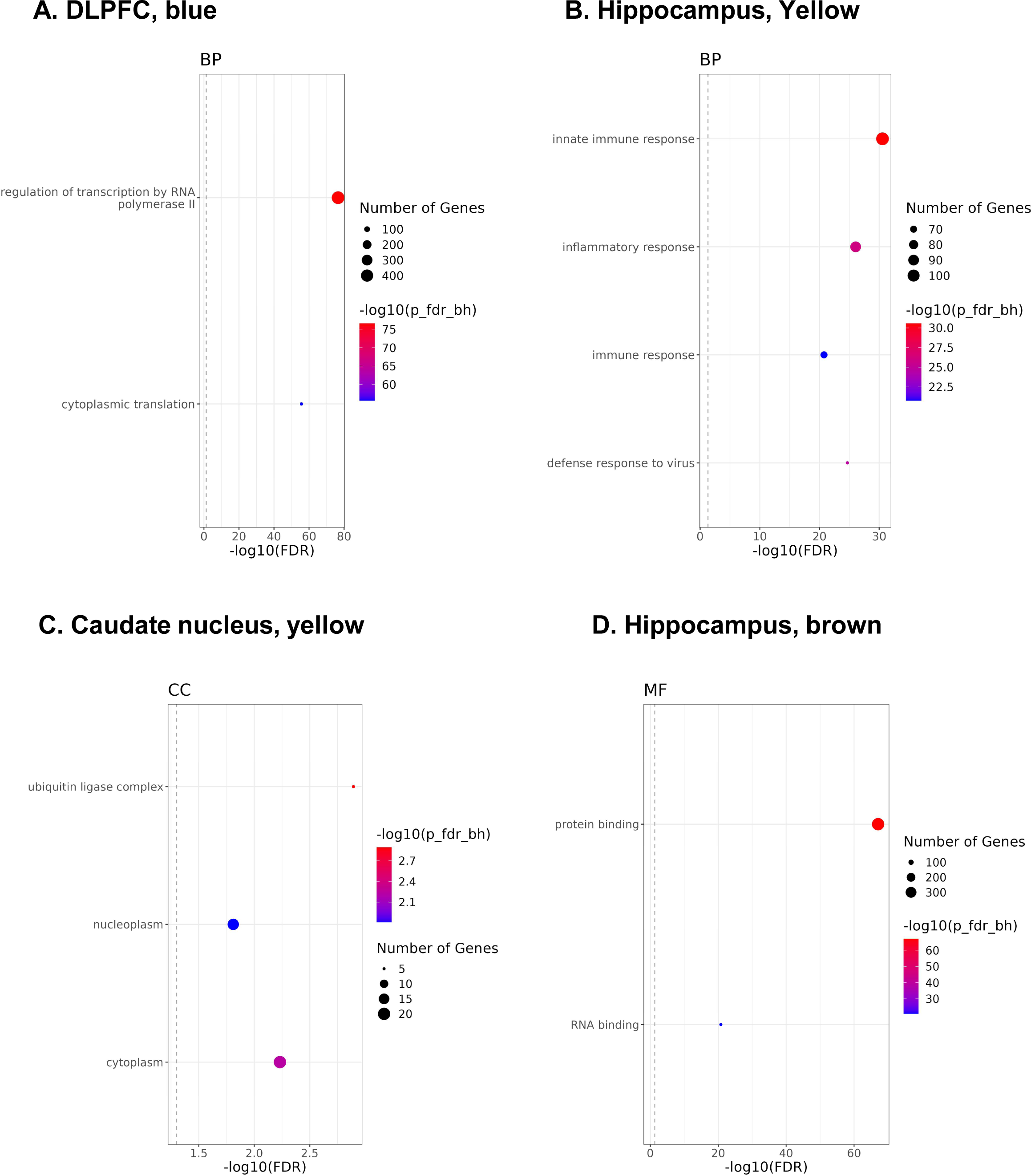
Gene ontology enrichment of gene– and repeat-dense WGCNA modules. A) Gene ontology enrichment of gene-dense DLPFC blue module B) Gene ontology enrichment of gene-dense hippocampus Yellow module C) Gene ontology enrichment of repeat-dense caudate nucleus Yellow module D) Gene ontology enrichment of repeat-dense hippocampus Brown module.

**Figure S4.**
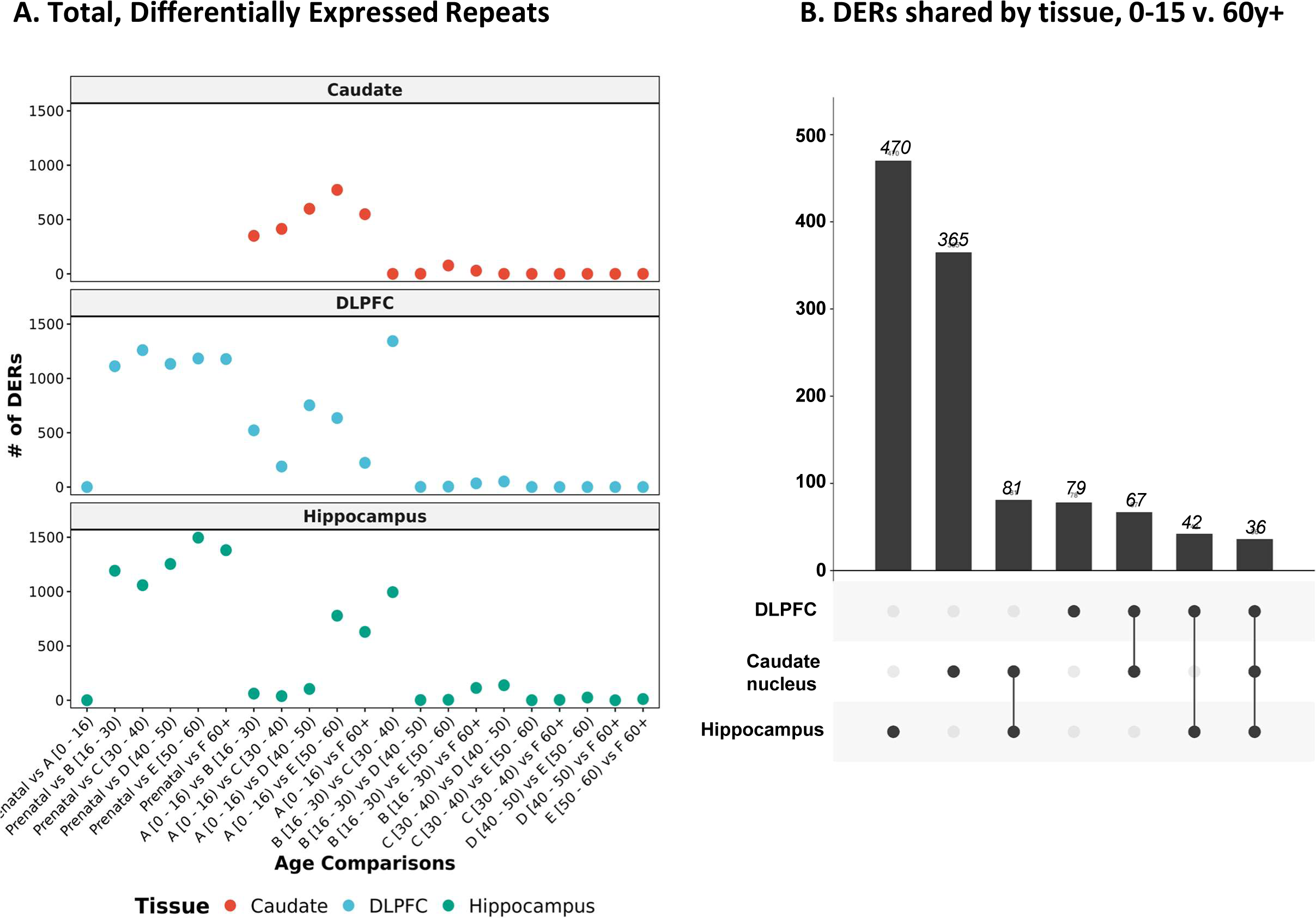
Distribution of differentially expressed repeats (DERs) across lifespan of neurotypical brain. A) Total differentially expressed repeats (DERs) across each age comparison in caudate nucleus, DLPFC, hippocampus. B) Upset Plot with the distribution of unique and shared 0-15 vs. 60y+ DERs by brain region.

**Figure S5.**
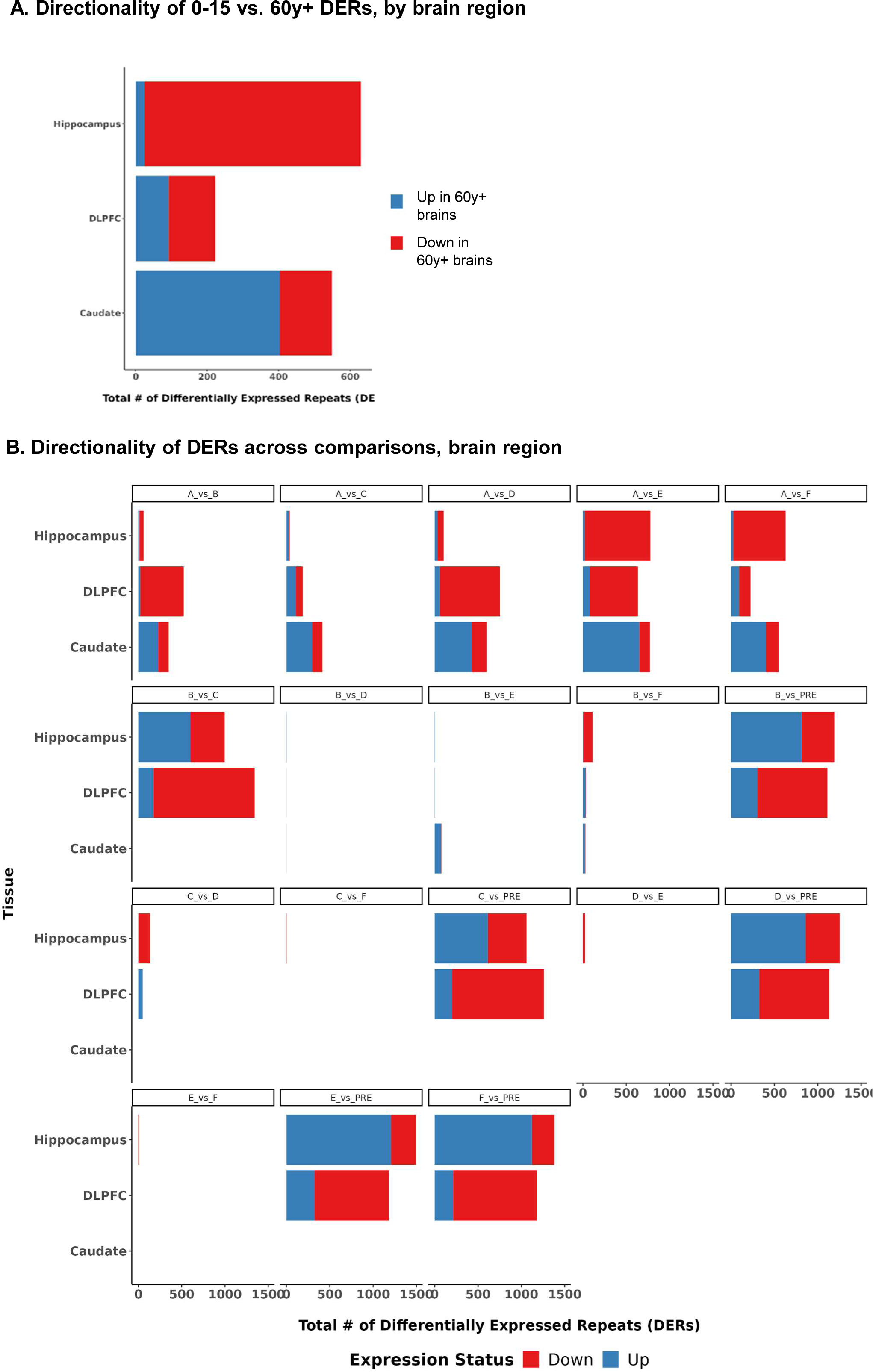
Directionality of DERs across lifespan of neurotypical brain. A) Up– and downregulated 0-15 vs. 60y+ DERs across each brain region. B) Up– and downregulated DERs across age comparisons by brain region.

**Figure S6.**
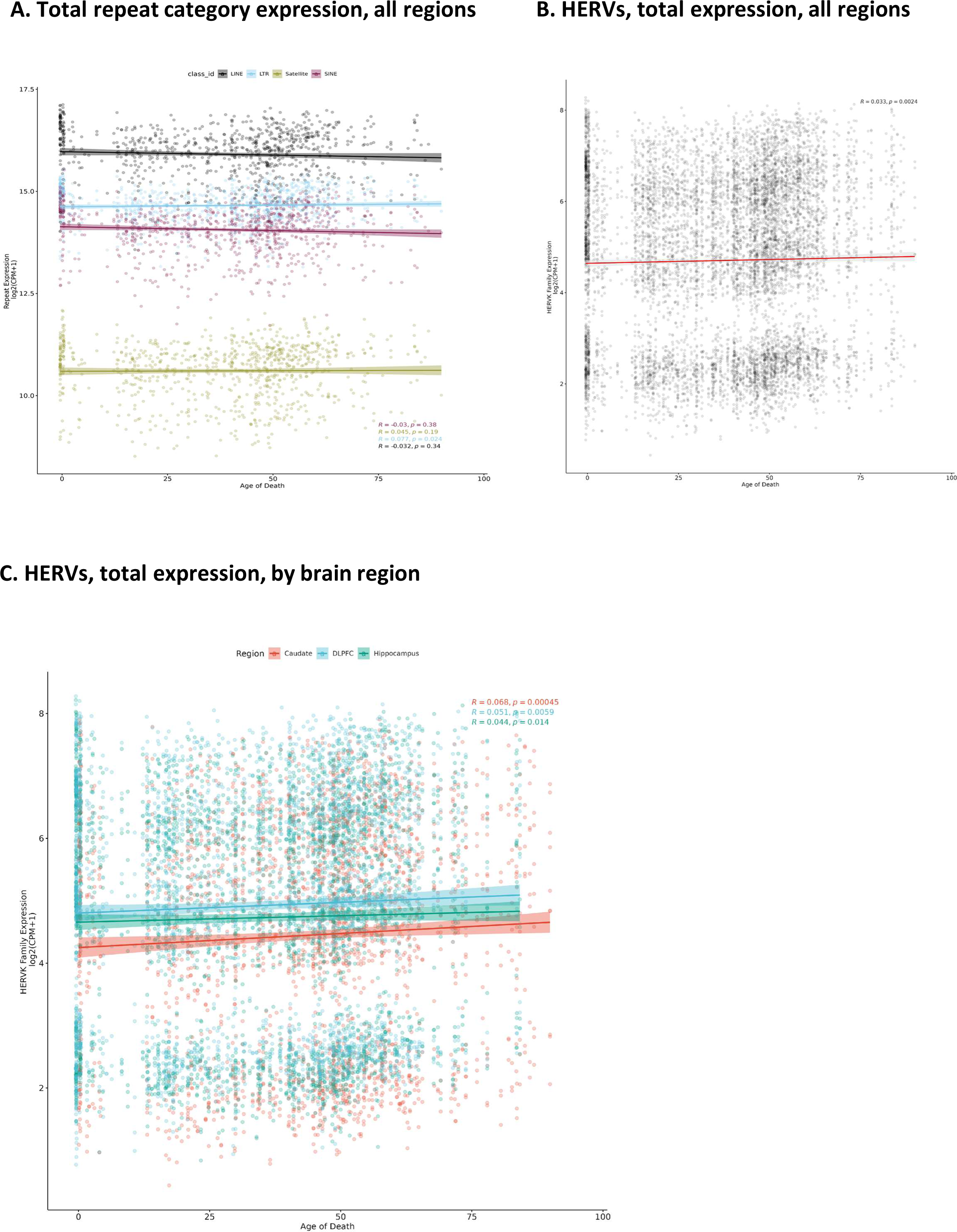
LTR and HERV-K expression correlates with age of death. A) Correlation of total LTR expression with age of death across all brain regions. B) Correlation of total HERV expression with age of death across all brain regions. C) Correlation of total HERV expression with age of death across each individual brain region.

## Supplemental Tables

**Table S1. Sample clinical data.** Information regarding all samples included in this project.

**Table S2. Scale-free topology fit index for each tissue-specific network (WGCNA).** Values obtained from pickSoftThreshold() function, to calculate the appropriate Beta of each region’s co-expression network (Caudate, DLPFC and Hippocampus).

**Table S3. WGCNA modules composition and features annotation for caudate nucleus, DLPFC, and hippocampus’ co-expression networks.** For the annotated genes, information regarding its chromosomal location is provided. For each repeats, information from its family and main class (obtained by RepeatMasker annotation) are also included. Features clustered in the ‘grey’ (null) module were excluded for further downstream analysis. Features without the module identification were absent for the WGCNA analysis of its respective co-expression network analysis.

**Table S4. Gene Ontology enrichment results for each WGCNA co-expression module for caudate nucleus, DLPFC and hippocampus co-expression networks.** Results were obtained with the GOATOOLS package. Only GO pathways with a FDR < 0.05 were considered as either enriched (’e’) or depleted (’p’). GO: Gene Ontology; NS: Gene Ontology Category; BP: Biological Process; MF: Molecular Function; CC: Cellular Component.

**Table S5. Power Analysis**. Power was derived from the sample sizes of each differential expression age group: Prenatal, A (0-15), B (16-29), C (30-39), D (40-49), E (50-59), F (60+), using TTestIndPower and FTestPower functions from Python’s statsmodels.stats.power.

**Table S6. Differential expression analysis applying limma-voom to features quantified by featureCounts algorithm.** Only features with a Benjamini-Hochberg false discovery rate (FDR) < 0.05 were considered as differentially expressed and included. AveExpr: average expression across all samples; logFC: estimate of the log2-fold-change corresponding to the effect; t: moderated t-statistic; B: log-odds that the gene is differentially expressed.

**Table S7. Differential expression analysis applying limma-voom and features quantified by TEcount algorithm.** Only features with a Benjamini-Hochberg false discovery rate < 0.05 were considered as differentially expressed and included. AveExpr: average expression across all samples; logFC: estimate of the log2-fold-change corresponding to the effect; t: moderated t-statistic; B: log-odds that the gene is differentially expressed.

**Table S8. Top intramodular hubs from each WGCNA module.** List of features (genes and/or repeats) that serve as intramodular hubs determined by the intramodularConnectivity.fromExpr() function. kTotal = total connectivity; kWhitin = intramodular connectivity; kOut = extra-modular connectivity; kDiff = the difference between the intra-modular and extra-modular connectivity.

## Data Availability

Raw data (.fastq files) are available under restricted access to protect research subjects. For total RNA data from prefrontal cortex and hippocampus, researchers can access the files via the Globus collections (jhpce#bsp2-dlpfc and jhpce#bsp2-hippo) at https://research.libd.org/globus/.

For caudate nucleus data, researchers can obtain access to FASTQ files via dbGaP accession phs003495.v1.p1 at https://www.ncbi.nlm.nih.gov/gap/.

## Acknowledgements

The authors gratefully acknowledge the families that have donated this tissue to the advancement of science. The authors would like to extend their appreciation to the Offices of the Chief Medical Examiner of Washington DC, Northern Virginia, Kalamazoo Michigan, Santa Clara County, University of North Dakota, and Maryland for the provision of brain tissue used in this work. The authors also extend their posthumous appreciation to to Dr. Llewellyn B. Bigelow and members of the LIBD Neuropathology Section for their work in assembling and curating the clinical and demographic information and organizing the Human Brain Tissue Repository of the Lieber Institute. We thank Dr. Alan Lorenzetti for the thoughtful feedback on this manuscript. We thank Johns Hopkins School of Medicine, Cellular and Molecular Medicine Graduate Program for additional support.

## Ethics Statement & Financial Disclosure

All sample identifiers used in this study are de-identified IDs and cannot reveal the identity of the study subjects. Additionally, all available sample clinical information also cannot reveal the identity of the study subjects. D.R.W. serves on the Scientific Advisory Boards of Sage Therapeutics and Pasithea Therapeutics. J.E.K. has served as a consultant for Merck on an antipsychotic drug trial. All other authors declare no competing interests. This work is supported by the Lieber Institute for Brain Development. JAE is supported by a NARSAD Young Investigator Grant from the Brain and Behavior Research Foundation and Collaborative Center For X-Linked Dystonia Parkinsonism and the MGH Collaborative Center for X-Linked Dystonia-Parkinsonism. JAE and ASF are supported by the Maryland Stem Cell Research Foundation. KJB is supported by T32 fellowship (T32MH015330) and K99 award (K99MD016964).

## Contributions

This work was collaborative, and many people contributed to many aspects of the project. Previously published postmortem sample collection and RNA sequencing were directed by Tom Hyde, Joel Kleinman and Daniel Weinberger at Lieber Institute for Brain Development. Repeat quantification was performed by Kynon Benjamin, Taylor Evans, Arthur Feltrin, Tarun Katipalli and Apua Paquola. WGCNA was largely conducted by Arthur Feltrin. Analysis and visualization of WGCNA results were largely conducted by Taylor Evans and Arthur Feltrin. Differential expression was performed by Kynon J Benjamin, Tarun Katipalli and Apua Paquola. Analysis and visualization of differential expression results was largely conducted by Taylor Evans with assistance from Kynon J Benjamin, Tarun Katipalli and Apua Paquola. The manuscript text and figures were largely performed by Taylor Evans, with assistance from Arthur Feltrin and Jennifer Erwin. Interpretation of findings was conducted by Taylor A. Evans with assistance from Arthur Feltrin, Apua Paquola, and Jennifer Erwin. The study was conceptualized and designed by Jennifer Erwin and Apua Paquola.

